# Importations of COVID-19 into African countries and risk of onward spread

**DOI:** 10.1101/2020.05.22.20110304

**Authors:** Haoyang Sun, Borame L Dickens, Alex R Cook, Hannah E Clapham

**Affiliations:** Saw Swee Hock School of Public Health, National University of Singapore, 12 Science Drive 2, Singapore 117549, Republic of Singapore

**Keywords:** Coronavirus, COVID-19, SARS-CoV-2, Africa, Mathematical modelling

## Abstract

**Background:** The emergence of a novel coronavirus (SARS-CoV-2) in Wuhan, China, at the end of 2019 has caused widespread transmission around the world. As new epicentres in Europe and America have arisen, of particular concern is the increased number of imported coronavirus disease 2019 (COVID-19) cases in Africa, where the impact of the pandemic could be more severe. We aim to estimate the number of COVID-19 cases imported from 12 major epicentres in Europe and America to each African country, as well as the probability of reaching 10,000 infections in total by the end of March, April, and May following viral introduction.

**Methods:** We used the reported number of cases imported from the 12 major epicentres in Europe and America to Singapore, as well as flight data, to estimate the number of imported cases in each African country. Under the assumption that Singapore has detected all the imported cases, the estimates for Africa were thus conservative. We then propagated the uncertainty in the imported case count estimates to simulate the onward spread of the virus, until 10,000 infections are reached or the end of May, whichever is earlier. Specifically, 1,000 simulations were run separately under two scenarios, where the reproduction number under the stay-at-home order was assumed to be 1.5 and 1.0 respectively.

**Findings:** We estimated Morocco, Algeria, South Africa, Egypt, Tunisia, and Nigeria as having the largest number of COVID-19 cases imported from the 12 major epicentres. Based on our 1,000 simulation runs, Morocco and Algeria’s estimated probability of reaching 10,000 infections by end of March was close to 100% under both scenarios. In particular, we identified countries with less than 100 cases in total reported by end of April whilst the estimated probability of reaching 10,000 infections by then was higher than 50% even under the more optimistic scenario.

**Conclusion:** Our study highlights particular countries that are likely to reach (or have reached) 10,000 infections far earlier than the reported data suggest, calling for the prioritization of resources to mitigate the further spread of the epidemic.

## Background

In late December 2019, a novel coronavirus (SARS-CoV-2) was identified among patients presenting with viral pneumonia in Wuhan city, China^1^. Since then the number of coronavirus disease 2019 (COVID-19) cases and deaths increased rapidly^2,3^, and the city was locked down by the Chinese government on 23^rd^ January 2020. By late February, there had only been limited importations from and to places outside China^4^. However, new epicentres in Europe and America emerged shortly thereafter, causing a second wave of importations that further accelerated the spread of the pandemic^4^. Most countries have since then imposed travel restrictions to prevent further importation of COVID-19 cases^5^. By 30^th^ April 2020, over three million cases and 200,000 deaths had been confirmed worldwide^4^.

A particular area of focus has been on countries in Africa, with worries about missed imported cases and what the impact will be of widespread transmission given the other heavy health burdens in these countries. The first confirmed case in Africa was reported in Egypt on 14^th^ February 2020, and two weeks later, the virus was found in sub-Saharan Africa with a reported case in Nigeria^4^. By the end of April, over 37,000 cases had been reported in the whole of Africa, with substantial variation in the reported cumulative incidence across different countries^4^. This inter-country heterogeneity can be due to a wide range of factors, such as the number of imported infections, the capacity to conduct tests for COVID-19, surveillance efforts, as well as travel and movement restrictions which vary widely from country to country depending on the local context^5^. The reported data alone thus do not provide a clear depiction of the outbreak situation especially in countries with very limited surveillance capacities, and additional studies are needed to narrow the knowledge gap between the reported data and the real disease burdens.

Previous work has estimated the risk of importation from China at the early stage of the pandemic^6^, assessed each African country’s capacity to respond to outbreaks^6^, systematically collated information on the importation events reported by the sub-Saharan countries^7^, and projected the spread of the epidemic seeded by the early cases represented in the World Health Organization Situation Reports^8^. It is still unclear how many infections may have been introduced to Africa from the new epicentres in Europe and America, although the reported case data do suggest that the size of this second wave of importations has been much larger than the first wave of importations from China^7^. In this study, we aim to estimate the number of COVID-19 cases imported from the major epicentres in Europe and America, and the magnitude of onward spread in each African country. This method is insensitive to the different testing and reporting systems that are in place in different countries.

## Methods

### Data

#### Case data

We collated data on the daily number of imported cases in Singapore reported by 31^st^ March from the following 12 epicentres: Austria, Belgium, France, Germany, Italy, Netherlands, Portugal, Spain, Switzerland, Turkey, United Kingdom, and United States, which accounted for over 90% of Singapore’s reported number of imported cases from countries outside of Asia^9^. These data will be used later to estimate the number of imported cases in Africa. In addition, we obtained the total number of cases (imported and autochthonous combined) reported by each African country by end of March and April from the World Health Organization’s situation reports^4^.

#### Government response data

For each country, we collated the date on which each of the following policies came into force: (1) banning non-citizens and non-residents from entry (the start date could vary depending on the epicentre country from which a visitor arrived); (2) mandatory (self-) quarantine for travellers arriving from each of the 12 epicentre countries mentioned earlier; (3) Stay-at-home order for all non-essential workers (hereinafter referred to as “stay-at-home order”). We reviewed the following sources: (1) country-level internal and international restrictions collated by the International SOS^5^, (2) Oxford COVID-19 Government Response Tracker^10^, (3) international travel restrictions collated by the International Air Transport Association^11^, as well as (4) Wikipedia, where a separate page was available for each country containing information regarding the government response. For each Wikipedia page, we manually reviewed the online reports listed in the references to exclude data with unconfirmed or unreliable sources. If stay-at-home order came into force in different states of the same country at different times, only the earliest date was recorded.

#### Travel data

We obtained the total number of air ticket bookings for each origin-destination route allowing for up to two connections during March 2017 from the Official Airline Guide. This will be used later to estimate the ratio of air passenger volumes between pairs of origin and destination countries, which we assumed to be relatively stable over time.

### Statistical analyses

#### Estimating the number of imported cases

For each African country *r*, we denote the daily number of air passengers that arrived from an epicentre country *e* by 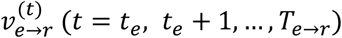, where *t_e_* refers to the start date of the COVID-19 epidemic in the epicentre country *e*, and *T_e→r_* refers to the last day that non-citizens and non-residents travelling from country *e* were allowed to enter country *r*. Each day the probability that an air passenger travelling from country *e* to country *r* was an imported case is denoted by 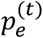, which we assume to be dependent on both the origin country *e* and time *t*, but independent from the destination country *r*. Hence, the total number of COVID-19 cases imported from an epicentre country *e* to an African country *r* by the time the travel ban came into force (denoted by *M_e→r_* below) can be approximated using a Poisson distribution (Refer to the supporting information for the derivation details):

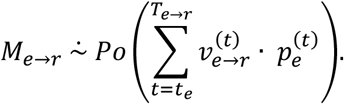

We used the imported COVID-19 case data reported by Singapore as well as flight data to provide a conservative estimate for *M_e_*_→_*_r_*, under the assumption that Singapore, being one of the countries with the highest surveillance capacity^12^, has detected all the imported cases. Owing to the delay from infection to hospital admission, we considered all cases imported from country *e* to Singapore that were *reported* by date *(T_e→r_ +* 9) (hereinafter denoted as *SG_e,r_*) based on Linton et al.’s estimated mean incubation period and time from illness onset to hospital admission^13^. We assumed that the ratio between the daily number of air travellers from epicentre *e* to country *r* and to Singapore remained stable in the presence of the changes in flight pattern in response to the COVID-19 pandemic. This allows us to model *M_e→r_* (and *SG_e,r_*) as Poisson random variables with mean parameters proportional to the numbers of air passengers travelling from epicentre *e* to country *r* (and to Singapore) using the March 2017 flight data (Refer to the supporting information for the derivation details):

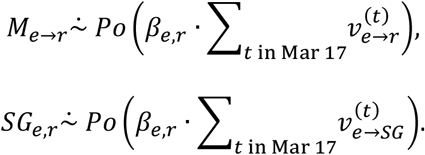

Here, *β_e,r_* refers to the proportionality constant to be estimated using the reported value of *SG_e,r_* and flight data, and was assigned a uniform prior with support (0, 1). We performed Markov Chain Monte Carlo to sample from the posterior distribution of *β_e,r_* using the JAGS software^14^, with 20,000 iterations burn-in and 150,000 iterations thinned for a posterior sample of size 5,000. The posterior sample for all the model parameters was then used to estimate the uncertainty distribution of the total number of COVID-19 cases imported from the 12 major epicentres to each country.

In March 2020, a spike in the number of cases imported from United Kingdom and United States was observed in Singapore, which was partly due to the increase in the number of returning Singaporean students studying overseas^15^. This change in flight patterns, however, may not be applicable to all African countries. Therefore, to be even more conservative, we also derived the imported case count estimates excluding United Kingdom and United States from the 12 epicentre countries previously considered. The resulting estimates were subsequently used in the simulations of the onward spread of SARS-CoV-2 to get our estimates of case numbers over time.

#### Simulating the onward transmission following importation

We performed 1,000 simulations drawing from our estimated distribution of the number of imported cases to project the onward spread of SARS-CoV-2 in each country up to 31^st^ May 2020 or the date when we estimate 10,000 infections was reached, whichever was earlier. The time of infection for the cases imported from country *e* to country *r* was simulated via resampling from the reporting dates of the *SG_e,r_* cases, which was then shifted backwards by 9 days to account for the delay from infection to hospital admission based on Linton et al.’s estimates^13^. To account for the effect of quarantine measures on the onward transmission, we only included the estimated imported cases who had acquired the infection prior to the mandatory quarantine of travellers coming into force, so that the estimation of local SARS-CoV-2 spread is conservative. For each country and each day, we followed Cori et al. and expressed the total infectiousness of the infected individuals as the weighted sum of the past incident infections^16^, where the weight parameters were derived from the cumulative distribution function of COVID-19’s serial interval based on Nishiura et al.’s estimate^17^. The number of secondary cases produced by each COVID-19 case, in the absence of stay-at-home order, was assumed to follow a negative binomial distribution with mean 2 and dispersion parameter 0.58^8^. Once the stay-at-home order came into force, we created two scenarios for the percentage reduction of the reproduction number: (1) 25% reduction, and (2) 50% reduction. To be conservative, we assumed that the stay-at-home order, once implemented, can be sustained up to the end date of our simulations. We ran the simulation algorithm following Churcher et al.^18^, and derived the estimated probability of reaching 10,000 infections by the end of March, April, and May respectively for each country. (Refer to the supporting information for the implementation details)

## Results

We estimated Morocco, Algeria, South Africa, Egypt, Tunisia, and Nigeria as having the largest number of COVID-19 cases imported from the 12 new epicentres in Europe and America (Table 1 and Figure 1). All of these countries had their lower bound estimate of the imported case count exceeding 100 (Table 1). By contrast, nine countries (e.g. Lesotho, Eswatini, and South Sudan) were found to have a very low risk of importation, with the upper bound estimate of the imported case count below 10 (Table 1). In a more conservative scenario where United Kingdom and United States were excluded from the list of epicentre countries, the estimated number of imported cases did not change drastically for most countries, albeit with some exceptions such as Kenya, whose estimate decreased from 97 (95% CI: 75–120) to 27 (95% CI: 16–41) (Table 1).

**Table 1:** Estimated number of COVID-19 cases (with 95% credible interval) imported from the 12 new epicentres in Europe and America (second column), and after excluding United Kingdom and United States from the list of epicentre countries (third column) to create a more conservative estimate (refer to Methods for more details).

| Country | Estimated imported case count from 12 epicentres | Estimated imported case count from 10 epicentres |
| --- | --- | --- |
| Algeria | 671 (489–891) | 630 (449–851) |
| Angola | 110 (48–227) | 95 (34–212) |
| Benin | 12 (6–20) | 10 (4–18) |
| Botswana | 4 (1–9) | 1 (0–4) |
| Burkina Faso | 15 (7–24) | 13 (6–22) |
| Burundi | 2 (0–7) | 2 (0–5) |
| Cabo Verde | 116 (83–173) | 55 (27–109) |
| Cameroon | 38 (25–54) | 29 (18–44) |
| Central African Republic | 3 (0–7) | 3 (0–7) |
| Chad | 3 (0–8) | 2 (0–6) |
| Comoros | 2 (0–6) | 2 (0–6) |
| Congo | 18 (9–29) | 14 (6–25) |
| Congo DRC | 22 (12–36) | 17 (8–30) |
| Côte d'Ivoire | 47 (29–68) | 39 (22–60) |
| Djibouti | 7 (2–13) | 5 (1–10) |
| Egypt | 287 (233–353) | 173 (125–231) |
| Equatorial Guinea | 12 (5–22) | 9 (3–18) |
| Eritrea | 3 (0–8) | 1 (0–4) |
| Eswatini | 0 (0–2) | 0 (0–1) |
| Ethiopia | 45 (32–61) | 20 (10–31) |
| Gabon | 16 (7–26) | 14 (6–24) |
| Gambia | 24 (14–35) | 7 (2–14) |
| Ghana | 77 (58–98) | 16 (8–26) |
| Guinea | 16 (8–25) | 13 (6–22) |
| Guinea-Bissau | 10 (3–24) | 10 (2–24) |
| Kenya | 97 (75–120) | 27 (16–41) |
| Lesotho | 0 (0–2) | 0 (0–1) |
| Liberia | 6 (2–12) | 2 (0–6) |
| Libya | 14 (4–36) | 13 (3–34) |
| Madagascar | 21 (11–34) | 19 (10–31) |
| Malawi | 7 (2–13) | 1 (0–4) |
| Mali | 23 (13–36) | 21 (11–33) |
| Mauritania | 6 (2–12) | 5 (1–11) |
| Mauritius | 122 (94–154) | 66 (44–93) |
| Mayotte | 6 (2–13) | 6 (2–13) |
| Morocco | 742 (575–959) | 555 (391–765) |
| Mozambique | 24 (11–46) | 19 (7–40) |
| Namibia | 10 (4–17) | 6 (2–12) |
| Niger | 8 (3–14) | 6 (2–13) |
| Nigeria | 160 (130–192) | 28 (17–41) |
| Rwanda | 12 (6–20) | 5 (1–11) |
| Réunion | 75 (46–114) | 74 (45–113) |
| Sao Tome and Principe | 8 (1–21) | 8 (1–20) |
| Senegal | 96 (69–129) | 83 (57–115) |
| Seychelles | 35 (23–49) | 22 (12–34) |
| Sierra Leone | 14 (7–22) | 2 (0–6) |
| Somalia | 9 (4–16) | 2 (0–6) |
| South Africa | 342 (287–404) | 119 (89–159) |
| South Sudan | 2 (0–6) | 1 (0–3) |
| Sudan | 23 (13–37) | 12 (5–24) |
| Tanzania | 58 (42–75) | 23 (13–36) |
| Togo | 11 (5–19) | 8 (3–15) |
| Tunisia | 214 (157–285) | 195 (137–264) |
| Uganda | 16 (8–25) | 7 (2–13) |
| Zambia | 15 (8–24) | 3 (0–7) |
| Zimbabwe | 30 (20–42) | 3 (0–7) |

**Figure 1:**
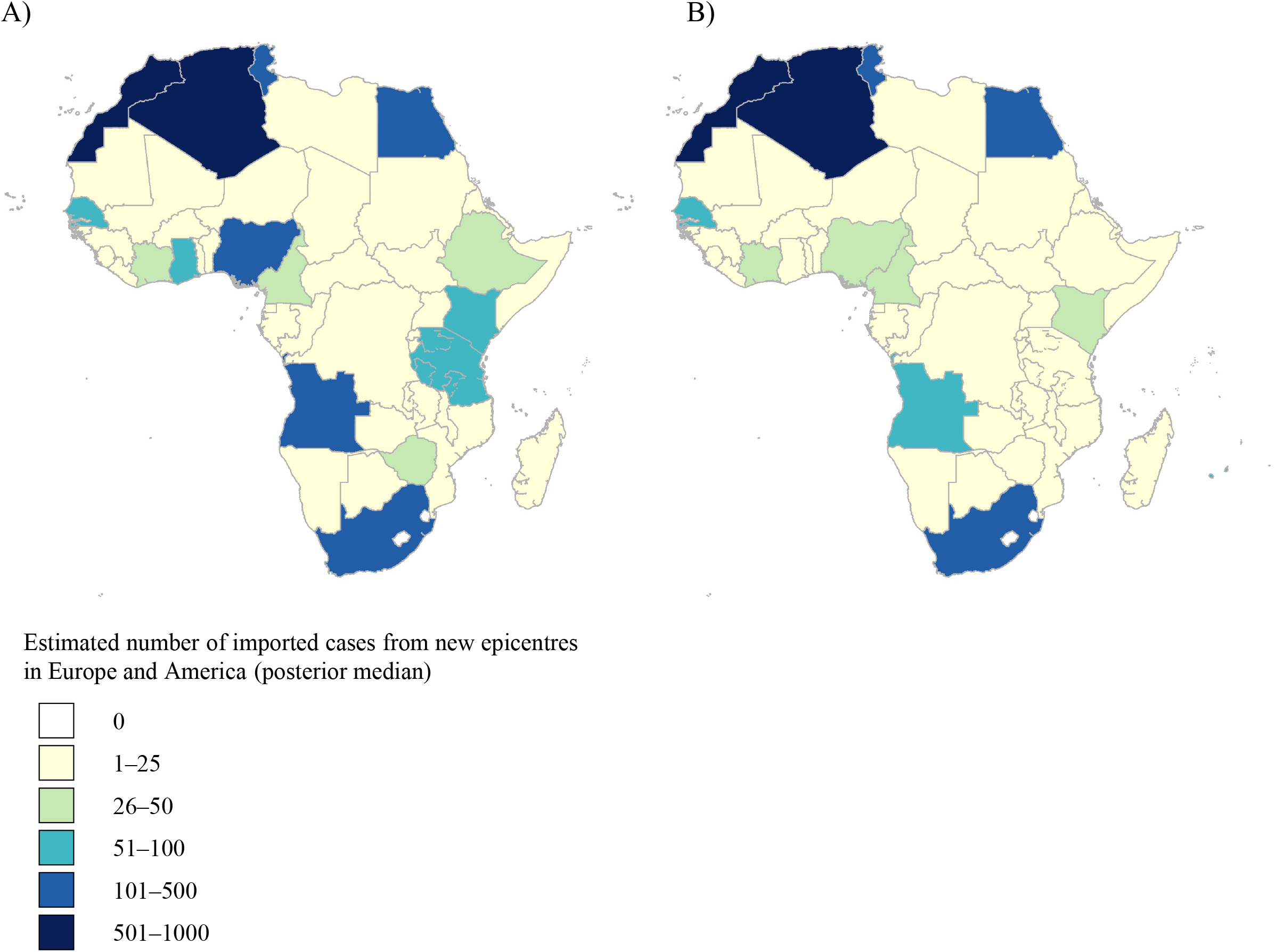
Posterior median estimates of the number of imported COVID-19 infections. Results include imported infections from (A) all the 12 major epicentres in Europe and America, and (B) 10 epicentres only, after excluding United Kingdom and United States to create a more conservative estimate (refer to Methods for more details).

Based on our 1,000 simulations of the onward SARS-CoV-2 spread, both Morocco and Algeria’s estimated probability of reaching 10,000 infections by end of March was close to 100% under both scenarios that we considered (Figures 2A, 2D), whilst the reported total number of cases in each country by end of March was ~500 (Figure 2G). Under the assumption that stay-at-home order reduces the reproduction number to 1.5, we found four African countries where the estimated probability of reaching 10,000 infections by end of March was higher than 50% (Figure 2A). This number quickly rose to 34 countries reaching this number of infections by the end of April, and 47 countries by end May (Figures 2B, 2C). For the alternative scenario where the reproduction number is reduced to 1.0 by stay-at-home order, the numbers of African countries with a higher-than-50% estimated probability of reaching 10,000 infections by end of March, April, and May were 3, 23, and 32 respectively (Figures 2D–2F). Notably, four countries (Angola, Gambia, Mozambique, and Sao Tome and Principe) were found to have reported less than 100 cases by end of April whilst the estimated probability of reaching 10,000 infections by then was higher than 50% even under the more optimistic scenario (Figures 2E, 2H), suggesting that a very substantial number of cases may have been undetected. The percentiles of the uncertainty distribution for the date by which 10,000 infections are reached in each country under the two scenarios were shown in Table 2.

**Figure 2:**
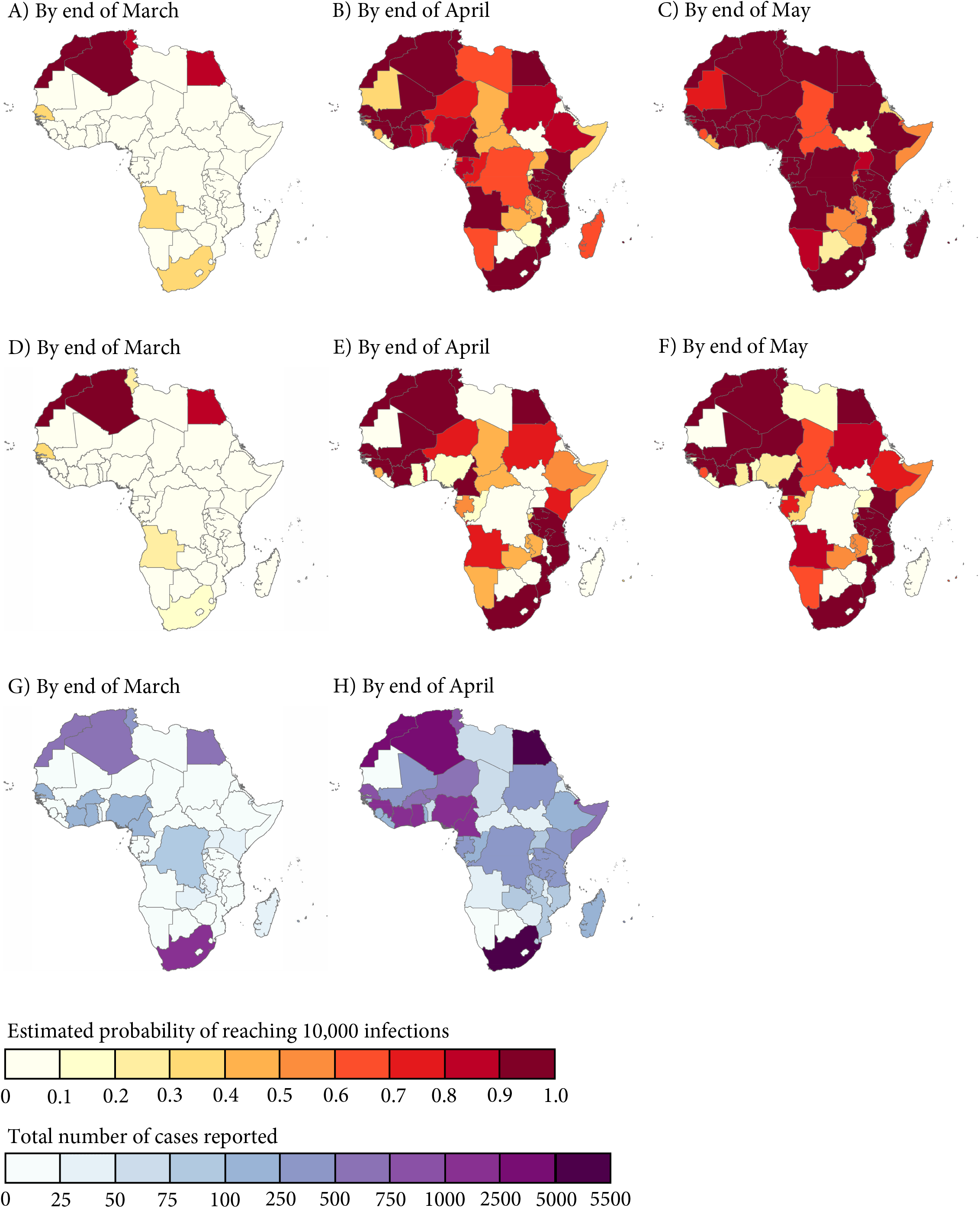
Estimated probability of reaching 10,000 infections as well as the reported total number of cases by each country. Stay-at-home order was assumed to reduce the reproduction number to (A–C) 1.5 and (D–F) 1.0 respectively. Reproduction number in the absence of stay-at-home order in each country was assumed to be 2. Reported total number of cases (G–H) were extracted from the World Health Organization’s situation reports.

**Table 2:**
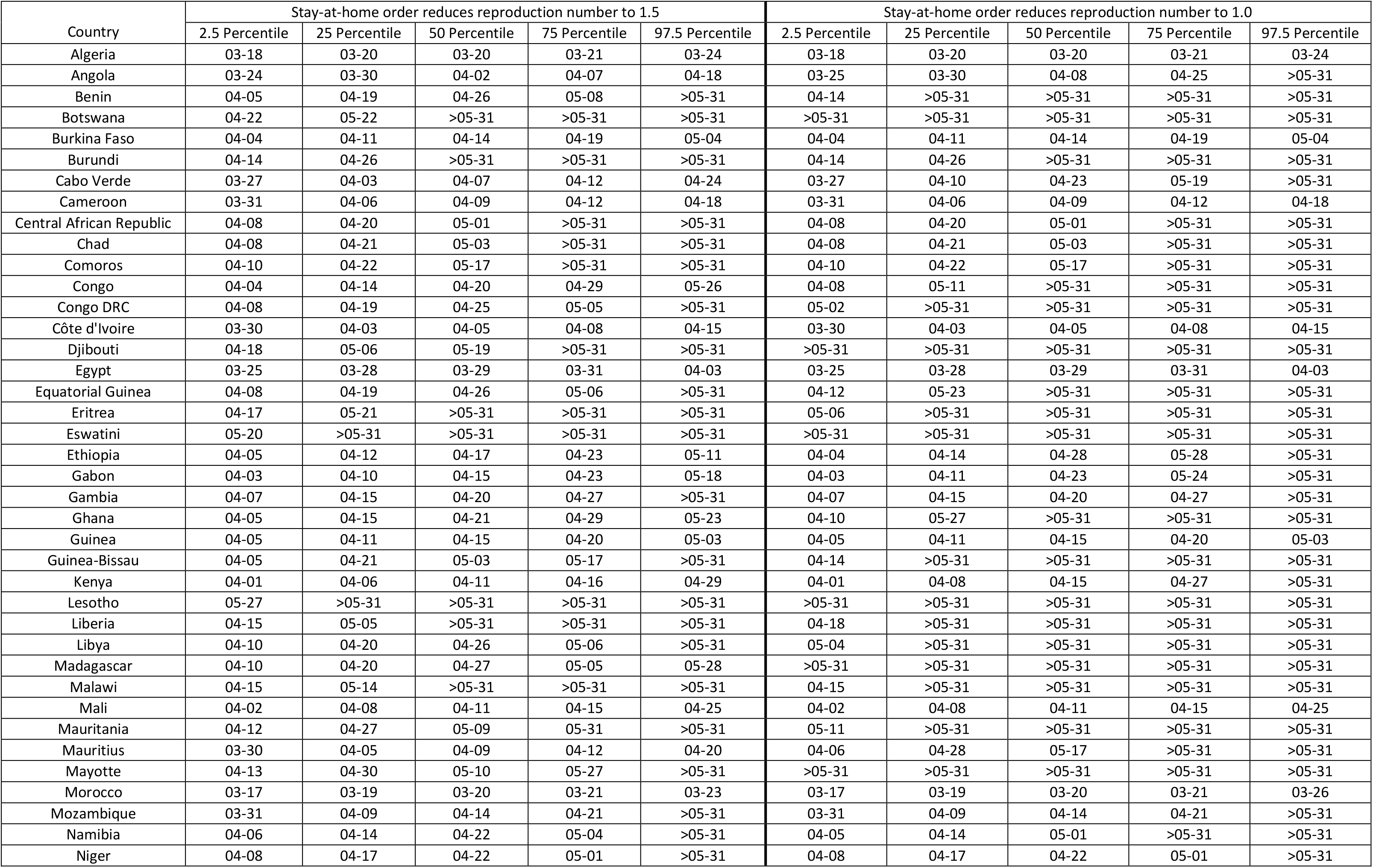
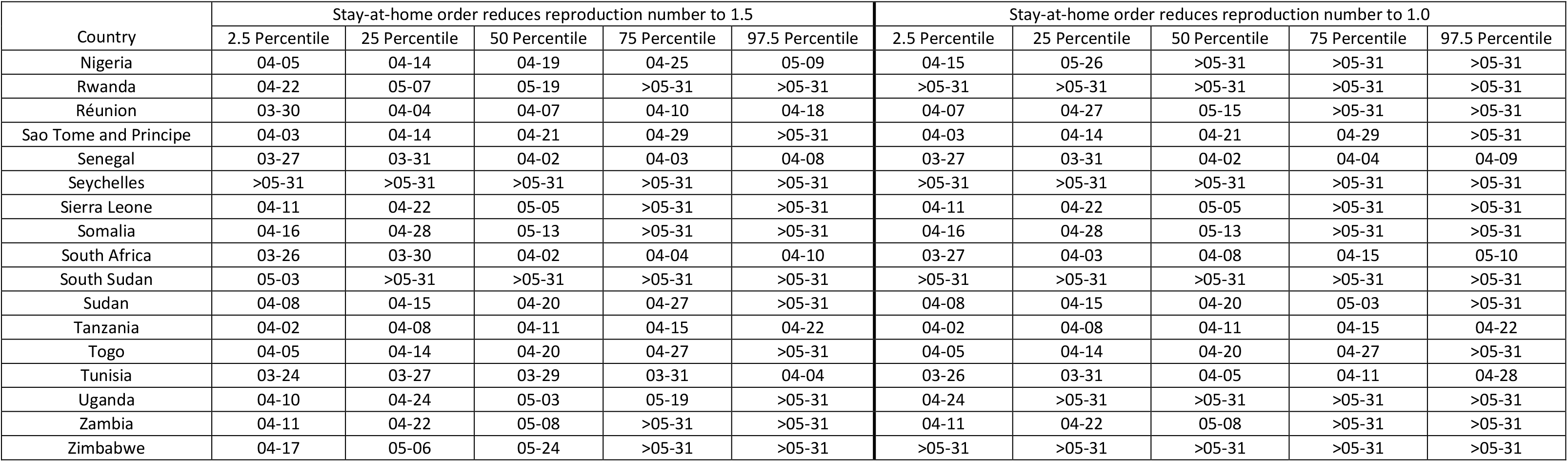
Summary statistics for the estimated date by which 10,000 infections are reached in each African country. Reproduction numbers used for the simulation were 2.0 before, and 1.5 or 1.0 after stay-at-home order came into force in each country. Simulations were performed until 31^st^ May, or 10,000 infections are reached, whichever is earlier, based on 1,000 model runs.

| Country | Stay-at-home order reduces reproduction number to 1.5 |  |  |  |  | Stay-at-home order reduces reproduction number to 1.0 |  |  |  |  |
| --- | --- | --- | --- | --- | --- | --- | --- | --- | --- | --- |
|  | 2.5 Percentile | 25 Percentile | 50 Percentile | 75 Percentile | 97.5 Percentile | 2.5 Percentile | 25 Percentile | 50 Percentile | 75 Percentile | 97.5 Percentile |
| Algeria | 03-18 | 03-20 | 03-20 | 03-21 | 03-24 | 03-18 | 03-20 | 03-20 | 03-21 | 03-24 |
| Angola | 03-24 | 03-30 | 04-02 | 04-07 | 04-18 | 03-25 | 03-30 | 04-08 | 04-25 | >05-31 |
| Benin | 04-05 | 04-19 | 04-26 | 05-08 | >05-31 | 04-14 | >05-31 | >05-31 | >05-31 | >05-31 |
| Botswana | 04-22 | 05-22 | >05-31 | >05-31 | >05-31 | >05-31 | >05-31 | >05-31 | >05-31 | >05-31 |
| Burkina Faso | 04-04 | 04-11 | 04-14 | 04-19 | 05-04 | 04-04 | 04-11 | 04-14 | 04-19 | 05-04 |
| Burundi | 04-14 | 04-26 | >05-31 | >05-31 | >05-31 | 04-14 | 04-26 | >05-31 | >05-31 | >05-31 |
| Cabo Verde | 03-27 | 04-03 | 04-07 | 04-12 | 04-24 | 03-27 | 04-10 | 04-23 | 05-19 | >05-31 |
| Cameroon | 03-31 | 04-06 | 04-09 | 04-12 | 04-18 | 03-31 | 04-06 | 04-09 | 04-12 | 04-18 |
| Central African Republic | 04-08 | 04-20 | 05-01 | >05-31 | >05-31 | 04-08 | 04-20 | 05-01 | >05-31 | >05-31 |
| Chad | 04-08 | 04-21 | 05-03 | >05-31 | >05-31 | 04-08 | 04-21 | 05-03 | >05-31 | >05-31 |
| Comoros | 04-10 | 04-22 | 05-17 | >05-31 | >05-31 | 04-10 | 04-22 | 05-17 | >05-31 | >05-31 |
| Congo | 04-04 | 04-14 | 04-20 | 04-29 | 05-26 | 04-08 | 05-11 | >05-31 | >05-31 | >05-31 |
| Congo DRC | 04-08 | 04-19 | 04-25 | 05-05 | >05-31 | 05-02 | >05-31 | >05-31 | >05-31 | >05-31 |
| Côte d'Ivoire | 03-30 | 04-03 | 04-05 | 04-08 | 04-15 | 03-30 | 04-03 | 04-05 | 04-08 | 04-15 |
| Djibouti | 04-18 | 05-06 | 05-19 | >05-31 | >05-31 | >05-31 | >05-31 | >05-31 | >05-31 | >05-31 |
| Egypt | 03-25 | 03-28 | 03-29 | 03-31 | 04-03 | 03-25 | 03-28 | 03-29 | 03-31 | 04-03 |
| Equatorial Guinea | 04-08 | 04-19 | 04-26 | 05-06 | >05-31 | 04-12 | 05-23 | >05-31 | >05-31 | >05-31 |
| Eritrea | 04-17 | 05-21 | >05-31 | >05-31 | >05-31 | 05-06 | >05-31 | >05-31 | >05-31 | >05-31 |
| Eswatini | 05-20 | >05-31 | >05-31 | >05-31 | >05-31 | >05-31 | >05-31 | >05-31 | >05-31 | >05-31 |
| Ethiopia | 04-05 | 04-12 | 04-17 | 04-23 | 05-11 | 04-04 | 04-14 | 04-28 | 05-28 | >05-31 |
| Gabon | 04-03 | 04-10 | 04-15 | 04-23 | 05-18 | 04-03 | 04-11 | 04-23 | 05-24 | >05-31 |
| Gambia | 04-07 | 04-15 | 04-20 | 04-27 | >05-31 | 04-07 | 04-15 | 04-20 | 04-27 | >05-31 |
| Ghana | 04-05 | 04-15 | 04-21 | 04-29 | 05-23 | 04-10 | 05-27 | >05-31 | >05-31 | >05-31 |
| Guinea | 04-05 | 04-11 | 04-15 | 04-20 | 05-03 | 04-05 | 04-11 | 04-15 | 04-20 | 05-03 |
| Guinea-Bissau | 04-05 | 04-21 | 05-03 | 05-17 | >05-31 | 04-14 | >05-31 | >05-31 | >05-31 | >05-31 |
| Kenya | 04-01 | 04-06 | 04-11 | 04-16 | 04-29 | 04-01 | 04-08 | 04-15 | 04-27 | >05-31 |
| Lesotho | 05-27 | >05-31 | >05-31 | >05-31 | >05-31 | >05-31 | >05-31 | >05-31 | >05-31 | >05-31 |
| Liberia | 04-15 | 05-05 | >05-31 | >05-31 | >05-31 | 04-18 | >05-31 | >05-31 | >05-31 | >05-31 |
| Libya | 04-10 | 04-20 | 04-26 | 05-06 | >05-31 | 05-04 | >05-31 | >05-31 | >05-31 | >05-31 |
| Madagascar | 04-10 | 04-20 | 04-27 | 05-05 | 05-28 | >05-31 | >05-31 | >05-31 | >05-31 | >05-31 |
| Malawi | 04-15 | 05-14 | >05-31 | >05-31 | >05-31 | 04-15 | >05-31 | >05-31 | >05-31 | >05-31 |
| Mali | 04-02 | 04-08 | 04-11 | 04-15 | 04-25 | 04-02 | 04-08 | 04-11 | 04-15 | 04-25 |
| Mauritania | 04-12 | 04-27 | 05-09 | 05-31 | >05-31 | 05-11 | >05-31 | >05-31 | >05-31 | >05-31 |
| Mauritius | 03-30 | 04-05 | 04-09 | 04-12 | 04-20 | 04-06 | 04-28 | 05-17 | >05-31 | >05-31 |
| Mayotte | 04-13 | 04-30 | 05-10 | 05-27 | >05-31 | >05-31 | >05-31 | >05-31 | >05-31 | >05-31 |
| Morocco | 03-17 | 03-19 | 03-20 | 03-21 | 03-23 | 03-17 | 03-19 | 03-20 | 03-21 | 03-26 |
| Mozambique | 03-31 | 04-09 | 04-14 | 04-21 | >05-31 | 03-31 | 04-09 | 04-14 | 04-21 | >05-31 |
| Namibia | 04-06 | 04-14 | 04-22 | 05-04 | >05-31 | 04-05 | 04-14 | 05-01 | >05-31 | >05-31 |
| Niger | 04-08 | 04-17 | 04-22 | 05-01 | >05-31 | 04-08 | 04-17 | 04-22 | 05-01 | >05-31 |

Table 2: Summary statistics for the estimated date by which 10,000 infections are reached in each African country.
| Country | Stay-at-home order reduces reproduction number to 1.5 |  |  |  |  | Stay-at-home order reduces reproduction number to 1.0 |  |  |  |  |
| --- | --- | --- | --- | --- | --- | --- | --- | --- | --- | --- |
|  | 2.5 Percentile | 25 Percentile | 50 Percentile | 75 Percentile | 97.5 Percentile | 2.5 Percentile | 25 Percentile | 50 Percentile | 75 Percentile | 97.5 Percentile |
| Nigeria | 04-05 | 04-14 | 04-19 | 04-25 | 05-09 | 04-15 | 05-26 | >05-31 | >05-31 | >05-31 |
| Rwanda | 04-22 | 05-07 | 05-19 | >05-31 | >05-31 | >05-31 | >05-31 | >05-31 | >05-31 | >05-31 |
| Réunion | 03-30 | 04-04 | 04-07 | 04-10 | 04-18 | 04-07 | 04-27 | 05-15 | >05-31 | >05-31 |
| Sao Tome and Principe | 04-03 | 04-14 | 04-21 | 04-29 | >05-31 | 04-03 | 04-14 | 04-21 | 04-29 | >05-31 |
| Senegal | 03-27 | 03-31 | 04-02 | 04-03 | 04-08 | 03-27 | 03-31 | 04-02 | 04-04 | 04-09 |
| Seychelles | >05-31 | >05-31 | >05-31 | >05-31 | >05-31 | >05-31 | >05-31 | >05-31 | >05-31 | >05-31 |
| Sierra Leone | 04-11 | 04-22 | 05-05 | >05-31 | >05-31 | 04-11 | 04-22 | 05-05 | >05-31 | >05-31 |
| Somalia | 04-16 | 04-28 | 05-13 | >05-31 | >05-31 | 04-16 | 04-28 | 05-13 | >05-31 | >05-31 |
| South Africa | 03-26 | 03-30 | 04-02 | 04-04 | 04-10 | 03-27 | 04-03 | 04-08 | 04-15 | 05-10 |
| South Sudan | 05-03 | >05-31 | >05-31 | >05-31 | >05-31 | >05-31 | >05-31 | >05-31 | >05-31 | >05-31 |
| Sudan | 04-08 | 04-15 | 04-20 | 04-27 | >05-31 | 04-08 | 04-15 | 04-20 | 05-03 | >05-31 |
| Tanzania | 04-02 | 04-08 | 04-11 | 04-15 | 04-22 | 04-02 | 04-08 | 04-11 | 04-15 | 04-22 |
| Togo | 04-05 | 04-14 | 04-20 | 04-27 | >05-31 | 04-05 | 04-14 | 04-20 | 04-27 | >05-31 |
| Tunisia | 03-24 | 03-27 | 03-29 | 03-31 | 04-04 | 03-26 | 03-31 | 04-05 | 04-11 | 04-28 |
| Uganda | 04-10 | 04-24 | 05-03 | 05-19 | >05-31 | 04-24 | >05-31 | >05-31 | >05-31 | >05-31 |
| Zambia | 04-11 | 04-22 | 05-08 | >05-31 | >05-31 | 04-11 | 04-22 | 05-08 | >05-31 | >05-31 |
| Zimbabwe | 04-17 | 05-06 | 05-24 | >05-31 | >05-31 | >05-31 | >05-31 | >05-31 | >05-31 | >05-31 |

## Discussion

Our study has estimated the size of the second wave of COVID-19 importations in each African country from the 12 major epicentres in Europe and America. This allows us to narrow the knowledge gap between the observed and actual number of importations, so that the unfolding of the epidemic seeded by the imported infections can be better projected especially in countries with very low testing capacities.

In the first wave of importations of infections from Wuhan, China, to other places outside China we estimated that most places at risk were in Asia, Europe and USA^19^. Though there were links between China and African countries, these were fewer than those between China and the rest of Asia, Europe and USA ^19^. The shut down in China severely curtailed continuing importations out of China and so these importations rapidly stopped.

Lower initial importations into Africa compared to Asia and Europe certainly tallies with what has been seen. There have been very few reported cases in Africa in the first wave of importations, and no reports of onward transmission. There was much discussion at the time whether the lack of reported imported cases in Africa was because imported cases were not being picked up. This may be some of the story, but our analysis would suggest that this was not the whole story, and it was more that the early risk of importation into Africa was lower than other places^19^. However the results we present in this paper estimate that this risk has dramatically increased with the spread of the virus in Europe and the USA. This also tallies with what we have seen, as countries in Africa started to report their first imported cases from Europe and the USA^4^. As of April 30^th^ 2020, South Africa had reported the highest number of cases at 5350^4^, and we estimated South Africa to have had one of the highest numbers of imported cases from the new epi-centres, although it was also rated highest at risk in Africa of importations from China in previous analysis^6^. Senegal is one of the countries for whom the risk has notably increased from the risk of importation from China as estimated in previous analyses^6,19^. We only considered importations from the major epicentres in Europe and America, and so the number of importations from all countries will be even higher.

Our study provides countries with information on the estimated timing of reaching 10,000 infections, which can be used for planning. Under the assumption that stay-at-home order reduces the reproduction number from 2.0 to 1.5, our estimates suggest that a number of African countries will reach (or have reached) 10,000 infections even earlier than the predictions of Pearson et al.^8^ This could be due to a number of imported infections being undetected and hence not reflected in the situation reports, as well as the delay from infection to reporting, both of which were accounted for in our study. Notably, we estimated two countries in North Africa, namely Algeria and Morocco, as having the highest probabilities of reaching 10,000 infections by the end of March, which may have occurred even prior to the lockdown. Countries in Sub-Saharan Africa having the earliest estimated timings of reaching 10,000 infections include Angola, Côte d’Ivoire, Senegal, and South Africa. In countries where stringent social distancing measures have yet to be implemented at the time of writing (e.g. Tanzania), the unfolding of the epidemic was estimated to be substantially faster than previous estimates suggest^8^. On the other hand, we projected that countries such as Seychelles will reach 10,000 infections later than Pearson et al.’s forecasts^8^ owing to the stay-at-home order. The epidemic was found to be further slowed down in many countries when we assumed the reproduction number to be reduced by 50% due to stay-at-home order.

Many countries in Africa have considerable experience in dealing with other infectious disease outbreaks, most notably Ebola, and will be able to call upon that experience for COVID-19. Countries hit in this third wave of transmission, including those in Africa have some advantage as there have been a variety of responses from around the world from which to assess what to do or not to do. However there will need to be consideration of how effective measures can be adapted to different settings^20^. Issues such as high HIV prevalence in some countries, and a younger demographic may both affect the cases and deaths observed in different ways. This relationship however is yet to be determined and there will need to be rapid research in countries in Africa to determine what the risk of disease is in different populations and how best to respond in light of many other competing health priorities.

Many countries in Africa are on high alert for incoming cases from Europe and USA, taking measures such as quarantine of arrivals or shutting down travel from affected countries. This is a sensible response given the vast amount of transmission on-going in these places. However as travel is either maintained or reopened between countries closer by, risk of importations from other countries should continue to be considered. Close attention should therefore be paid to where will be the next epicentre, perhaps within Africa, and how this could translate into imported cases for each country, particularly for those countries that we estimate to have experienced lower numbers of imported cases previously and therefore lower onward transmission.

Not accounted for in our study currently is the impact of less stringent interventions on the local SARS-CoV-2 spread, such as the effect of prohibiting large public gatherings, closure of social venues and schools, and restrictions on inter-district travels. It is still unclear as to whether and to what extent these interventions were effective in their local context, and hence in our simulations we only considered stay-at-home order for all non-essential workers as an effective intervention to reduce local transmission. Future modelling work considering the impact of different interventions in different places will be vital for determining how each country can continue to respond.

In addition, we have made simplifying assumptions about the change in travel patterns in response to the pandemic in each African country relative to that in Singapore, due to the unavailability of 2020 flight data. Despite these limitations, most of our model assumptions throughout the analyses have been fairly conservative to avoid inflating the projections of the SARS-CoV-2 spread. For example, the reported number of imported cases in Singapore was assumed to be complete, and the risk of returning citizens carrying SARS-CoV-2 after travel restrictions came into force in each African country was also not included. Simulations of the onward spread of the virus were based on the estimated number of imported infections from the selected 10 epicentre countries, and stay-at-home order was assumed to be effective (reproduction numbers being 1.5 and 1.0 in the two scenarios we considered) and sustainable. In light of these conservative assumptions, any countries found to have a high probability of reaching 10,000 infections by end March or April—especially those with very limited cases detected—need urgent actions.

## Conclusions

In conclusion, our study provides model estimates of the number of COVID-19 cases imported from major epicentres in Europe and America to each country in Africa, as well as simulation results of the onward epidemic spread. Our results highlight particular countries that are likely to reach (or have reached) 10,000 infections far earlier than the reported data suggest, calling for the prioritization of resources to mitigate the further spread of the epidemic.

## Data Availability

Data on the reported number of imported cases in Singapore are available from https://www.moh.gov.sg/covid-19/past-updates
Government response data used in this study have been included within the Additional file 1.
Supporting information has been included within the Additional file 2.

## List of abbreviations

COVID-19: Coronavirus disease 2019
SARS-CoV-2: Severe acute respiratory syndrome coronavirus 2

## Declarations

### Ethics approval and consent to participate

Not applicable.

### Consent for publication

Not applicable.

### Availability of data and materials

Data on the reported number of imported cases in Singapore are available from https://www.moh.gov.sg/covid-19/past-updates

Flight data used in this study were purchased from the Official Airline Guide.

Government response data used in this study have been included within the Additional file 1.

Supporting information has been included within the Additional file 2.

### Authors’ contributions

HS and HC designed the study and HS carried out the analysis. All authors contributed to the results interpretation, writing of the manuscript and approved it before submission.

## Acknowledgements

Not applicable.

